# Prevalence and Trends in Cardiovascular Risk Factors Among Middle Aged Persons from Five Race and Ethnic Groups in the United States: A Longitudinal Analysis of Two Cohort Studies

**DOI:** 10.1101/2024.09.27.24314520

**Authors:** Havisha Pedamallu, Zeynab Aghabazaz, Nicola Lancki, Luis A. Rodriguez, Juned Siddique, Meena Moorthy, Nilay S. Shah, Norrina B. Allen, Alka M. Kanaya, Namratha R. Kandula

**Author notes:** These authors contributed equally to this work (Havisha Pedamallu, Zeynab Aghabazaz). **Corresponding Author:** Namratha R. Kandula, MD, MPH 675 N St Clair St Ste 18-200, Chicago, IL 60611.

## Abstract

**Importance:** It is well established that people of South Asian background have a high burden of atherosclerotic cardiovascular disease (ASCVD). However, few studies have comprehensively examined if South Asian adults in the United States (US) develop cardiovascular risk factors at younger ages than adults from other race and ethnic groups.

**Objective:** To compare the prevalence and change in ASCVD risk factors across age strata by race and ethnic group.

**Design:** We combined data from the Mediators of Atherosclerosis in South Asians Living in America (MASALA) and the Multi-Ethnic Study of Atherosclerosis (MESA) cohort studies. Longitudinal data from all eligible participants at all available exam visits were used to estimate the prevalence of risk factors at ages 45 and 55 years for each race and ethnic group.

**Setting:** Multicenter longitudinal cohort study in 7 field centers across the U.S.

**Participants:** The baseline study sample included individuals free of clinical ASCVD; 554 South Asians, 796 White, 588 Black, 517 Hispanic/Latino, and 245 Chinese adults aged 45-55 years were included.

**Exposures:** Self-identified race (Black, Chinese, South Asian, or White) or ethnic group (Hispanic/Latino).

**Main Outcome(s) and Measure(s):** Prevalence of clinical (prediabetes & diabetes, hypertension, dyslipidemia, BMI) and behavioral (diet quality, alcohol use, exercise) ASCVD risk factors at age 45 and age 55.

**Results:** At age 45, South Asian men and women had the highest prevalence of pre-diabetes and diabetes and higher prevalence of hypertension compared to White, Chinese, and Hispanic/Latino men and women. South Asian men had a higher prevalence of dyslipidemia than White, Chinese, and Black men, and South Asian women had a higher prevalence than Chinese and Black women. All groups had worse diet quality than South Asian men and women at age 45, and most also had higher rates of alcohol use.

**Conclusions and Relevance:** We observed significant differences in the prevalence of risk factors for South Asian adults compared to adults from other U.S. race and ethnic groups at age 45 years. Understanding trends and disparities in cardiovascular risk and protective factors across the life course can help equitably improve prevention and treatment strategies for US populations.

**Key Points:** *Question:* Do South Asian adults have a higher burden of cardiovascular risk factors at age 45 years compared adults from other race and ethnic groups?

*Findings:* In this study of 2754 adults from two cohort studies, the prevalence of prediabetes and diabetes at age 45 years was higher among South Asians than in Black, Chinese, Hispanic and White adults; hypertension prevalence was higher among South Asians than all groups except Black adults.

*Meaning:* South Asian adults have a higher prevalence of several clinical cardiovascular risk factors at a younger age.

## INTRODUCTION

South Asian individuals (people who trace their ancestry from Bangladesh, Bhutan, India, Maldives, Nepal, Pakistan, and Sri Lanka) have high rates of atherosclerotic cardiovascular disease (ASCVD), with estimates approximating rates 2.5 times higher compared to non-Hispanic White and East Asian populations^1^. On average, South Asians develop coronary artery disease (CAD) up to a decade earlier than the average age at which CAD develops in most people in the United States (US) ^2^. A quarter of heart attacks occur under age 40 for South Asians and 50% occur under age 50. In clinical guidelines, South Asian ancestry is considered a “risk enhancing factor”^3^.

While South Asian in the US have an overall higher prevalence of ASCVD compared to non-Hispanic White US populations that manifests at younger ages, there is limited research to know if the prevalence of ASCVD risk factors at younger ages is higher among South Asian populations compared to other race and ethnic groups. Therefore, the objective of this study was to combine longitudinal data from the Mediators of Atherosclerosis in South Asians Living in America (MASALA) study and the Multiethnic Study of Atherosclerosis (MESA) to estimate the prevalence of cardiovascular risk factors during middle age, and to test whether the prevalence of these risk factors and their change over time differ between South Asians compared to Black, Chinese, Hispanic/Latino and White adults. If South Asians develop ASCVD risk factors at younger ages than other race and ethnic groups, this finding could lead to earlier screening, prevention, and treatment for this at-risk group.

## METHODS

### Study Population

This study was conducted using longitudinal data from all available exams of the Mediators of Atherosclerosis in South Asians Living in America (MASALA) and the Multi-Ethnic Study of Atherosclerosis (MESA) studies, which are both multicenter prospective cohort studies examining the development and determinants of clinical ASCVD risk factors, subclinical ASCVD, and ASCVD outcomes.

The study design for MASALA has been published elsewhere^4^. Briefly, MASALA included 1164 South Asian (defined as those with Indian, Pakistani, Bangladeshi, Nepali, and Sri Lankan ancestry) participants who were CVD free between 40 to 84 years of age recruited through two field centers in San Francisco, CA and Chicago, IL; Data were collected at baseline (Exam 1) for 906 participants in 2010-2013 with follow up Exam 2 data collection in 2016-2018 for 748 participants. An additional 258 participants were recruited in 2017-2018 and their baseline (Exam 1A) data are included.

The study design for MESA has also been published ^5^. Briefly, MESA included 6,814 White, Black, Hispanic/Latino, and Chinese men and women free from clinical CVD between 45-84 years of age. Baseline exam 1 data were collected between 2000-2002 with repeat examinations every 2 years. There have been 6 exams, the most recent being 2016-2018. Participants were recruited through 6 field centers: Forsyth County, NC; Northern Manhattan and the Bronx, NY; Baltimore City and Baltimore County, MD; St. Paul, MN; Chicago and the village of Maywood, IL; and Los Angeles County, CA.

### Study Sample and Exclusion Criteria

The analysis was restricted to participants between ages 45 to 55 at the baseline exam to minimize bias related to the cohorts’ inclusion criteria of being free from ASCVD at baseline. It is likely that the older individuals enrolled in MASALA and MESA may not be representative of the general older population since they were required to be free of clinical symptoms of ASCVD and ASCVD is more common at older ages. Therefore, we included 554 South Asians from the MASALA study and 796 White, 588 Black, 517 Hispanic/Latino, and 245 Chinese adults from the MESA study. Harmonized data from MASALA Exams 1/1A and 2 and MESA Exams 1 through 6 were combined. On average, our data set consisted of 3.3 exams per MESA participant and 1.4 exams per MASALA participant.

### Primary Exposure

The primary exposure was self-reported race (Black, Chinese, South Asian, or White) or ethnic group (Hispanic/Latino).

### Primary Outcome

The primary outcomes were clinical and behavioral ASCVD risk factors. These risk factors were chosen based on prior research establishing important risk factors in the development of ASCVD and the American Heart Association’s Life’s Essential 8^6^. Risk factor definitions were previously harmonized between the two data sets using a standardized set of definitions^7–9^. All risk factors were available at all exams, except diet and exercise at MESA exam 4. Diet was assessed for 87% of MESA participants at Exam 1, 72% of MESA participants at exam 5, 99% of MASALA participants at Exam 1, and 71% of MASALA participants at Exam 2.

Clinical risk factors include hypertension, pre-diabetes or diabetes status, dyslipidemia, and body mass index (BMI). Hypertension was defined as having a systolic blood pressure ≥140 mmHg, diastolic blood pressure ≥90 mmHg, or use of any antihypertensive medication according to guidelines in use at the time of the baseline exam^10^. Prediabetes was defined as having a fasting plasma glucose level between 100 and 125 mg/dL. Diabetes status was defined by self-reported use of glucose-lowering medications or fasting plasma glucose ≥ 126 mg/dL^11^. Dyslipidemia was determined using standard lipid level cut points (total cholesterol ≥ 200 mg/dL, low-density lipoprotein ≥160 mg/dL, triglycerides ≥150 mg/dL, or high-density lipoprotein < 40 mg/dL) using blood samples or self-report of currently taking lipid lowering therapy-statin/fibrate/niacin^12^. BMI was calculated using weight and height measurements at each exam visit and was treated as a continuous variable in the analysis. BMI at Exam 1 was reported as a categorical variable using race and ethnic group-specific cut-points (Chinese and South Asian Individuals: underweight BMI < 18.5, normal BMI 18.5 to 22.9, overweight 23 to 24.9, and obese BMI ≥ 25; All others: underweight BMI < 18.5, normal BMI 18.5 to 24.9, overweight 25 to 29.9, and obese BMI ≥ 30)^13, 14^.

Behavioral risk factors included alcohol use, physical activity level, and diet quality. Alcohol use was defined as currently drinking 1 or more drinks per week. Participants reported exercise using the Typical Week’s Physical Activity Questionnaire which was quantified as MET minutes/week^15^. Diet quality was determined by calculating the Alternative Health Eating Index-2010 (AHEI) score^9^. This is a dietary quality index that predicts the risk of chronic disease based on nutrient and food intake, with a range between 0 (worst) to 110 (best) points^9^.

### Statistical Analysis

Baseline characteristics (age, smoking status, education, insurance status, BMI) were calculated for each race and ethnic group. Comparisons across groups were tested using one-way ANOVAs for continuous variables and chi-squared tests of independence for categorical variables.

Our estimands of interest are the risk factor prevalences (for binary outcomes) or means (for continuous outcomes) at age 45 and their change over 10 years for each race and ethnic group and their differences between race and ethnic groups.

Using data from all eligible MESA and MASALA participants, we modeled clinical and behavioral risk factors as a function of age using mixed-effects linear regression models for continuous outcomes (BMI, physical activity, diet quality) and mixed-effects logistic regression models for binary outcomes (all other outcomes). Smoking status was not modeled as a longitudinal risk factor because it tended not to vary over time and cannot be modeled at age 45 and 55. All models controlled for baseline education and baseline insurance status and were fit separately by risk factor and sex.

To estimate risk factor prevalences at age 45 and their 10-year change, we used the method of marginal predicted means^16^ - the average value of the outcome averaging over all covariates—to estimate the prevalence of each risk factor at ages 45 and 55 for each race and ethnic group. These marginal predicted means were obtained from our mixed-effects models that use data from all eligible MESA and MASALA participants.

To compare risk factor prevalence between South Asians and the other race and ethnic groups, we first performed—separately by risk factor—an overall test at age 45 of whether the South Asian prevalence was significantly different from any of the other race and ethnic groups. If that test was significant, we subsequently performed the four pairwise tests comparing South Asians to each of the other race and ethnic groups.

To test if the change in risk factors between ages 45 and 55 differed for South Asians compared to the other groups, we calculated the difference (slope) between marginal means at ages 45 and 55 (i.e 55 minus 45) for all five race and ethnic groups. Then, we again performed an overall test of whether the South Asian slope was significantly different from the slopes of any of the other race and ethnic groups. If this test was significant, we then performed the four pairwise tests comparing the South Asian slope to the slope of each race and ethnic group. More information on estimation of risk factor prevalences, their change over time, and their comparison across race and ethnic groups in included in the Appendix. Since this is an exploratory analysis of observational data, we did not correct for multiple comparisons. All analyses and post-estimation processing were performed using STATA version 18.0^17^.

## RESULTS

### Baseline Characteristics

The average age of South Asian participants was 48.5 years at the baseline exam which was similar when compared to the other race and ethnic groups (Table 1). South Asian participants overall had higher education levels compared to the other race and ethnic groups. A majority (60.8%) of South Asians had more than a college level education compared to 32.9% of White participants, the next highest educated group. South Asians also had the highest never smoking prevalence (85.2%) at baseline with Chinese adults having a slightly lower prevalence (76.7%). Hispanic/Latino (53.0%), Black (48.0%), and White (45.5%) had much lower never smoking prevalences in comparison. A higher proportion of South Asian (86.8%), White (91.7%), and Black (81.0%) participants had private insurance compared to Chinese (68.2%) and Hispanic/Latino (67.3%) participants. When using race and ethnic group-specific BMI cut points, 20.9% of South Asian, 34.6% of White, 35.5% of Chinese, 15.8% of Black, and 16.1% of Latino participants had an underweight or normal BMI,

**Table 1:**
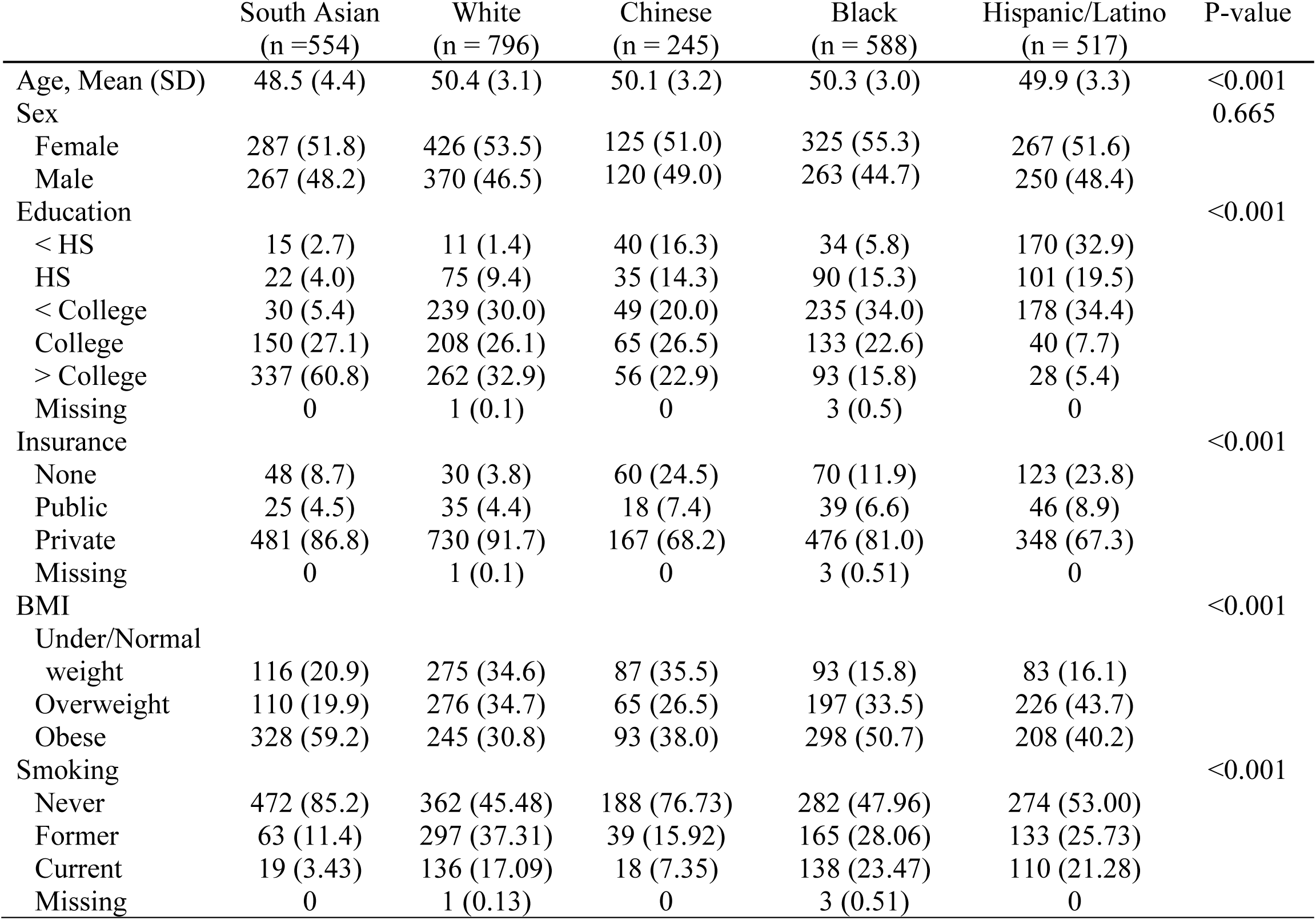
Baseline Characteristics at Exam 1, MASALA and MESA studies. Values are N (%) unless otherwise reported.

|  | South Asian<br>(n =554) | White<br>(n = 796) | Chinese<br>(n = 245) | Black<br>(n = 588) | Hispanic/Latino<br>(n = 517) | P-value |
| --- | --- | --- | --- | --- | --- | --- |
| Age, Mean (SD) | 48.5 (4.4) | 50.4 (3.1) | 50.1 (3.2) | 50.3 (3.0) | 49.9 (3.3) | <0.001 |
| Sex |  |  |  |  |  | 0.665 |
| Female | 287 (51.8) | 426 (53.5) | 125 (51.0) | 325 (55.3) | 267 (51.6) |  |
| Male | 267 (48.2) | 370 (46.5) | 120 (49.0) | 263 (44.7) | 250 (48.4) |  |
| Education |  |  |  |  |  | <0.001 |
| < HS | 15 (2.7) | 11 (1.4) | 40 (16.3) | 34 (5.8) | 170 (32.9) |  |
| HS | 22 (4.0) | 75 (9.4) | 35 (14.3) | 90 (15.3) | 101 (19.5) |  |
| < College | 30 (5.4) | 239 (30.0) | 49 (20.0) | 235 (34.0) | 178 (34.4) |  |
| College | 150 (27.1) | 208 (26.1) | 65 (26.5) | 133 (22.6) | 40 (7.7) |  |
| > College | 337 (60.8) | 262 (32.9) | 56 (22.9) | 93 (15.8) | 28 (5.4) |  |
| Missing | 0 | 1 (0.1) | 0 | 3 (0.5) | 0 |  |
| Insurance |  |  |  |  |  | <0.001 |
| None | 48 (8.7) | 30 (3.8) | 60 (24.5) | 70 (11.9) | 123 (23.8) |  |
| Public | 25 (4.5) | 35 (4.4) | 18 (7.4) | 39 (6.6) | 46 (8.9) |  |
| Private | 481 (86.8) | 730 (91.7) | 167 (68.2) | 476 (81.0) | 348 (67.3) |  |
| Missing | 0 | 1 (0.1) | 0 | 3 (0.51) | 0 |  |
| BMI |  |  |  |  |  | <0.001 |
| Under/Normal weight | 116 (20.9) | 275 (34.6) | 87 (35.5) | 93 (15.8) | 83 (16.1) |  |
| Overweight | 110 (19.9) | 276 (34.7) | 65 (26.5) | 197 (33.5) | 226 (43.7) |  |
| Obese | 328 (59.2) | 245 (30.8) | 93 (38.0) | 298 (50.7) | 208 (40.2) |  |
| Smoking |  |  |  |  |  | <0.001 |
| Never | 472 (85.2) | 362 (45.48) | 188 (76.73) | 282 (47.96) | 274 (53.00) |  |
| Former | 63 (11.4) | 297 (37.31) | 39 (15.92) | 165 (28.06) | 133 (25.73) |  |
| Current | 19 (3.43) | 136 (17.09) | 18 (7.35) | 138 (23.47) | 110 (21.28) |  |
| Missing | 0 | 1 (0.13) | 0 | 3 (0.51) | 0 |  |

### Risk Factor Prevalence at Age 45 among Men

South Asian men had the highest prevalence of pre-diabetes and diabetes (41.8%; Table 2) when compared to men of all other groups (White: 7.2%, Chinese: 18.9%, Black: 16.0%, Hispanic/Latino: 14.7%; p < 0.001). They had a significantly greater prevalence of hypertension compared to Chinese and Hispanic/Latino men and a significantly greater prevalence of dyslipidemia compared to Black men. They also had a significantly lower BMI than White, Black, and Hispanic/Latino men. South Asian men had significantly higher diet quality scores compared to men of all other race and ethnic groups (Table 4). They also had significantly lower alcohol use than Chinese men and higher level of exercise than White and Chinese men.

**Table 2:**
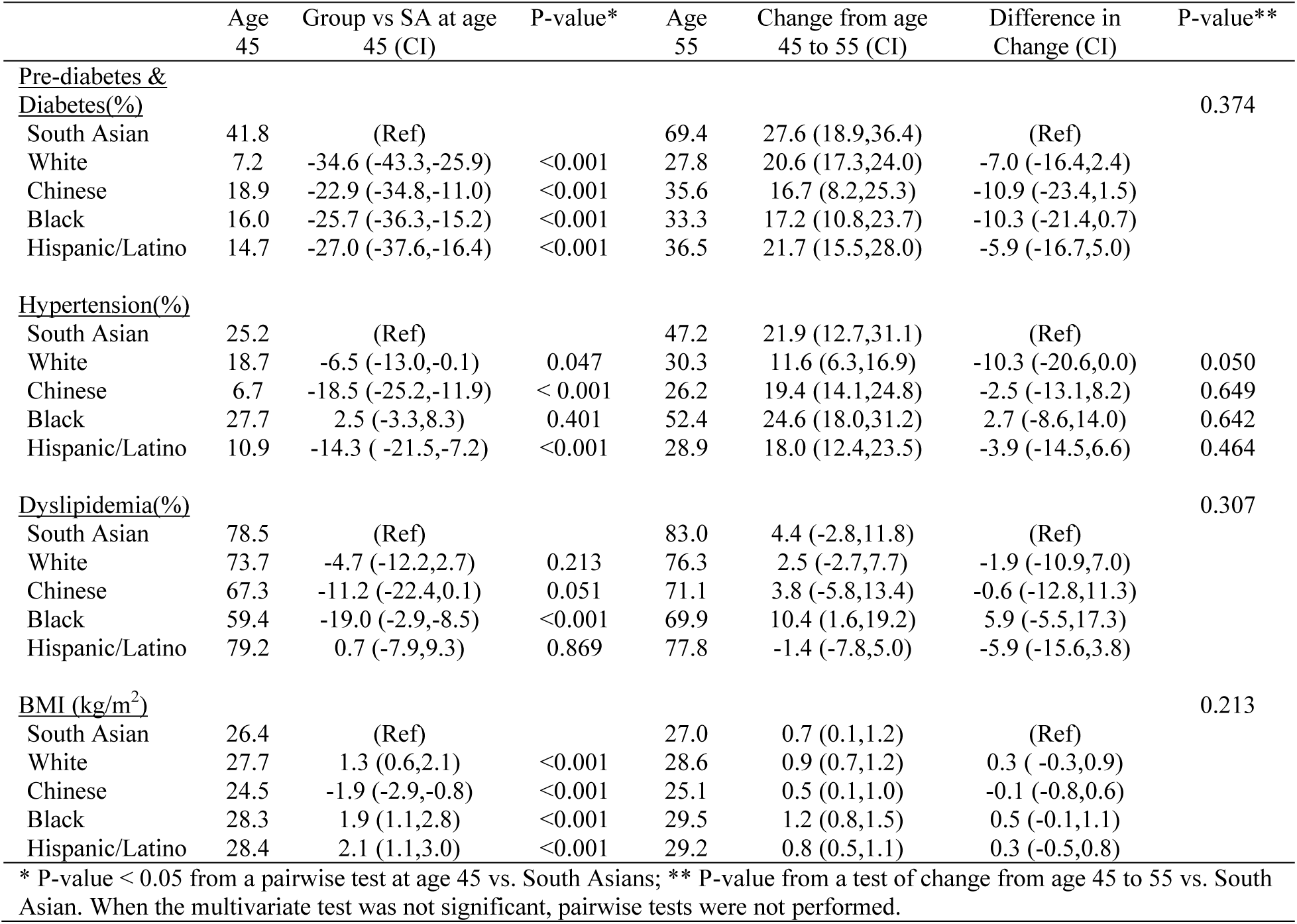
Clinical Risk Factors Stratified by Age and Race and Ethnic Group in Men, MASALA vs. MESA Groups.

**Table 3:**
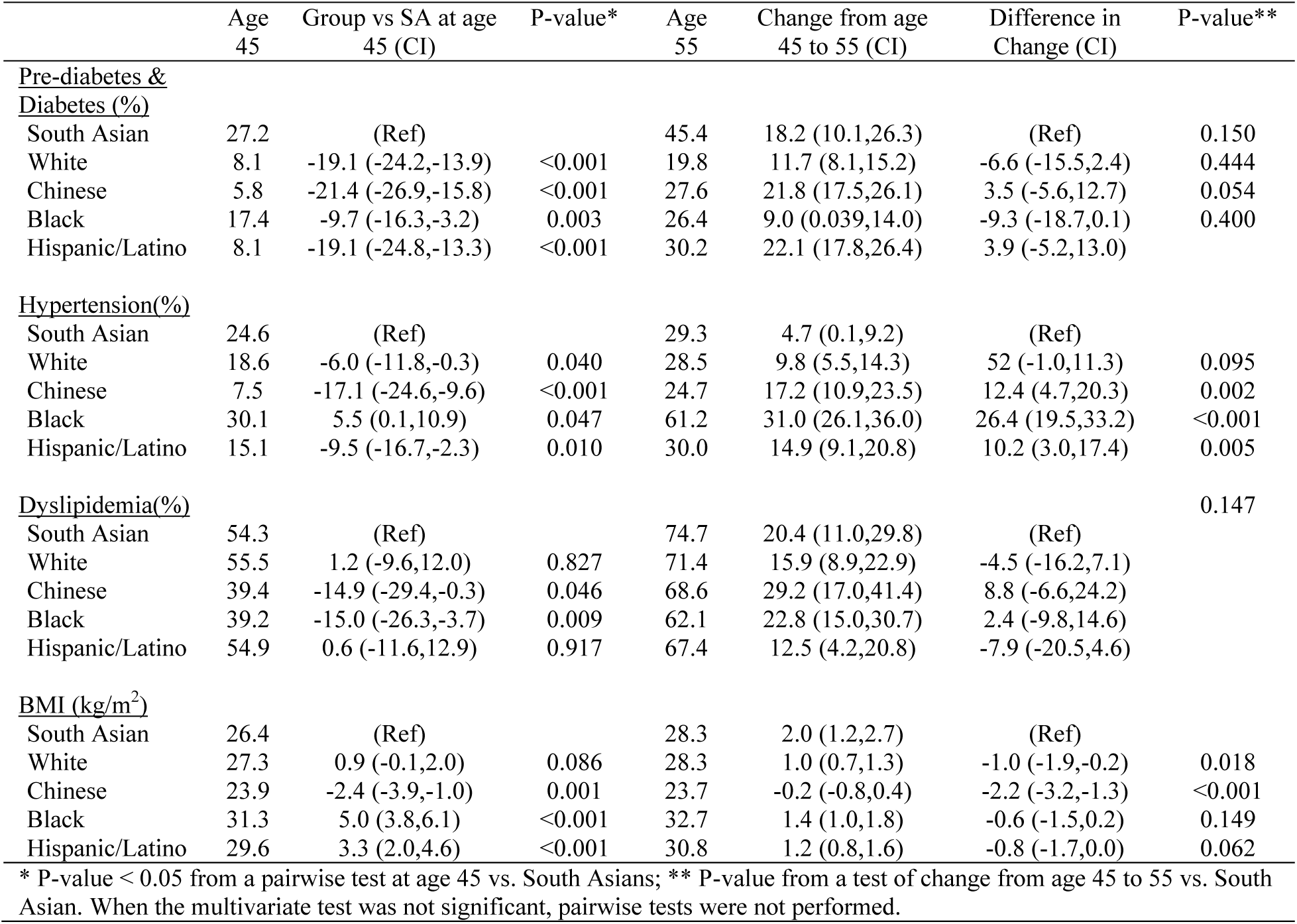
Clinical Risk Factors Stratified by Age and Race and Ethnic Group in Women, MASALA vs. MESA Groups.

**Table 4:**
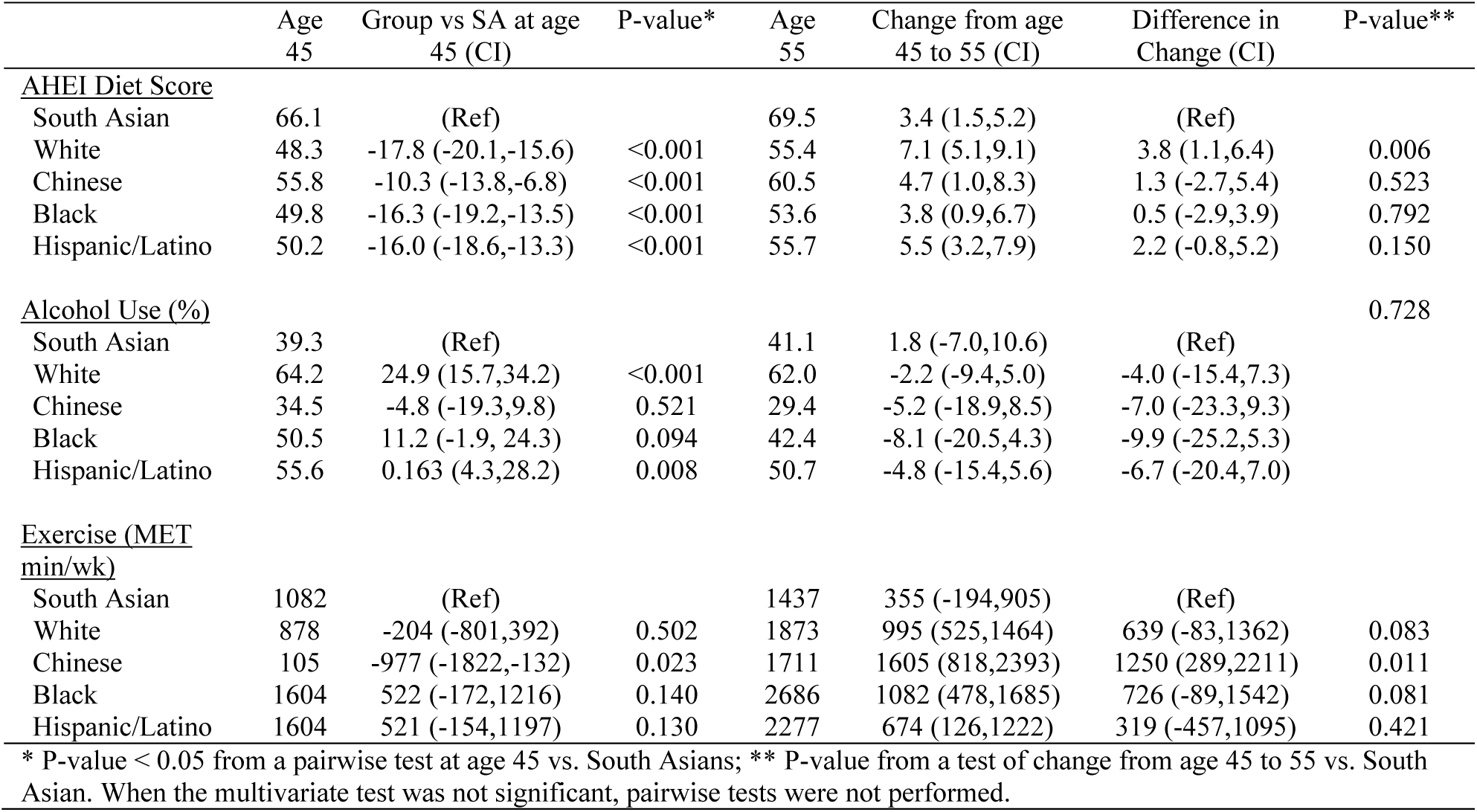
Behavioral Risk Factors Stratified by Age and Race and Ethnic Group in Men, MASALA vs. MESA Groups.

|  | Age<br>45 | Group vs SA at age<br>45 (CI) | P-value* | Age<br>55 | Change from age<br>45 to 55 (CI) | Difference in<br>Change (CI) | P-value** |
| --- | --- | --- | --- | --- | --- | --- | --- |
| <u>AHEI Diet Score</u> |  |  |  |  |  |  |  |
| South Asian | 66.1 | (Ref) |  | 69.5 | 3.4 (1.5,5.2) | (Ref) |  |
| White | 48.3 | -17.8 (-20.1,-15.6) | <0.001 | 55.4 | 7.1 (5.1,9.1) | 3.8 (1.1,6.4) | 0.006 |
| Chinese | 55.8 | -10.3 (-13.8,-6.8) | <0.001 | 60.5 | 4.7 (1.0,8.3) | 1.3 (-2.7,5.4) | 0.523 |
| Black | 49.8 | -16.3 (-19.2,-13.5) | <0.001 | 53.6 | 3.8 (0.9,6.7) | 0.5 (-2.9,3.9) | 0.792 |
| Hispanic/Latino | 50.2 | -16.0 (-18.6,-13.3) | <0.001 | 55.7 | 5.5 (3.2,7.9) | 2.2 (-0.8,5.2) | 0.150 |
| <u>Alcohol Use (%)</u> |  |  |  |  |  |  |  |
| South Asian | 39.3 | (Ref) |  | 41.1 | 1.8 (-7.0,10.6) | (Ref) | 0.728 |
| White | 64.2 | 24.9 (15.7,34.2) | <0.001 | 62.0 | -2.2 (-9.4,5.0) | -4.0 (-15.4,7.3) |  |
| Chinese | 34.5 | -4.8 (-19.3,9.8) | 0.521 | 29.4 | -5.2 (-18.9,8.5) | -7.0 (-23.3,9.3) |  |
| Black | 50.5 | 11.2 (-1.9, 24.3) | 0.094 | 42.4 | -8.1 (-20.5,4.3) | -9.9 (-25.2,5.3) |  |
| Hispanic/Latino | 55.6 | 0.163 (4.3,28.2) | 0.008 | 50.7 | -4.8 (-15.4,5.6) | -6.7 (-20.4,7.0) |  |
| <u>Exercise (MET</u><br><u>min/wk)</u> |  |  |  |  |  |  |  |
| South Asian | 1082 | (Ref) |  | 1437 | 355 (-194,905) | (Ref) |  |
| White | 878 | -204 (-801,392) | 0.502 | 1873 | 995 (525,1464) | 639 (-83,1362) | 0.083 |
| Chinese | 105 | -977 (-1822,-132) | 0.023 | 1711 | 1605 (818,2393) | 1250 (289,2211) | 0.011 |
| Black | 1604 | 522 (-172,1216) | 0.140 | 2686 | 1082 (478,1685) | 726 (-89,1542) | 0.081 |
| Hispanic/Latino | 1604 | 521 (-154,1197) | 0.130 | 2277 | 674 (126,1222) | 319 (-457,1095) | 0.421 |

\* P-value < 0.05 from a pairwise test at age 45 vs. South Asians; \*\* P-value from a test of change from age 45 to 55 vs. South Asian. When the multivariate test was not significant, pairwise tests were not performed.

**Table 5:** Behavioral Risk Factors Stratified by Age and Race and Ethnic Group in Women, MASALA vs. MESA groups.

|  | Age<br>45 | Group vs SA at age<br>45 (CI) | P-value* | Age<br>55 | Change from age<br>45 to 55 (CI) | Difference in<br>Change (CI) | P-value** |
| --- | --- | --- | --- | --- | --- | --- | --- |
| <u>AHEI Diet Score</u> |  |  |  |  |  |  |  |
| South Asian | 68.5 | (Ref) |  | 71.8 | 3.35 (1.5,5.2) | (Ref) |  |
| White | 51.5 | -17 (-19.2,-14.7) | <0.001 | 58.5 | 7.02 (5.2,8.9) | 3.67 (1.0,6.3) | 0.006 |
| Chinese | 58.6 | -9.8 (-13.1,-6.6) | <0.001 | 64.6 | 5.94 (2.7,9.2) | 2.59 (-1.2,6.3) | 0.176 |
| Black | 47.7 | -21 (-23.3,-18.2) | <0.001 | 53.5 | 5.78 (3.5,8.1) | 2.43 (-0.5,5.4) | 0.106 |
| Hispanic/Latino | 50.3 | -18.2 (-20.8,-15.6) | <0.001 | 56.6 | 6.32 (4.1,8.5) | 2.97 (0.1,5.9) | 0.045 |
| <u>Alcohol Use (%)</u> |  |  |  |  |  |  |  |
| South Asian | 18.6 | (Ref) |  | 17.4 | -1.2 (-7.0,4.6) | (Ref) | 0.051 |
| White | 42.8 | 24.1 (16.0,32.4) | <0.001 | 46.7 | 3.8 (-3.8,11.5) | 5.0 (-4.5,14.6) |  |
| Chinese | 25.8 | 0.072 (-4.4,18.7) | 0.226 | 14.8 | -11.0 (-23.4,1.4) | -9.8 (-23.5,3.9) |  |
| Black | 30.7 | 0.120 (4.1,20.0) | 0.003 | 21.4 | -9.2 (-16.4, -2.1) | -8.1 (-17.3,1.1) |  |
| Hispanic/Latino | 32.4 | 0.137 (4.8,22.7) | 0.002 | 23.8 | -8.6 (-16.5, -0.7) | -7.4 (-17.2,2.3) |  |
| <u>Exercise (MET</u><br><u>min/wk)</u> |  |  |  |  |  |  |  |
| South Asian | 1226 | (Ref) |  | 1429 | 203 (-263, 670) | (Ref) |  |
| White | 989 | -237 (-718, 243) | 0.334 | 1597 | 608 (242, 974) | 405 (-188, 998) | 0.181 |
| Chinese | 407 | -819 (-1503, -136) | 0.019 | 1267 | 861 (204, 1518) | 657 (-148, 1463) | 0.110 |
| Black | 1063 | -163 (-689, 362) | 0.542 | 2025 | 962 (525, 1400) | 759 (119, 1399) | 0.020 |
| Hispanic/Latino | 950 | -276 (-815, 264) | 0.316 | 1609 | 659 (219, 1098) | 455 (-185, 1096) | 0.164 |

\* P-value < 0.05 from a pairwise test at age 45 vs. South Asians; \*\* P-value from a test of change from age 45 to 55 vs. South Asian. When the multivariate test was not significant, pairwise tests were not performed.

### Trends Across Age Among Men

There were no significantly different changes in prevalence of clinical risk factors between age 45 and 55 for South Asian men versus men from the other groups, with the exception of White men having a smaller increase in the prevalence of hypertension between age 45 and 55 (21.9% for South Asian men, 11.6% for White men, p = 0.05). Among the behavioral risk factors, White men also had a larger improvement in AHEI diet score (7.1) than South Asian men (3.4, p = 0.006) over time. Chinese men had a larger increase in exercise (1605 MET min/wk) than South Asian men (355 MET min/wk, p = 0.011).

### Risk Factor Prevalence at Age 45 among Women

Similar to South Asian men, South Asian women had the highest prevalence of pre-diabetes and diabetes of women of all race and ethnic groups (Table 3). South Asian women also had the highest hypertension prevalence, and a significantly higher prevalence of dyslipidemia compared to Chinese and Black women. BMI among South Asian women was significantly higher than BMI in Chinese women and significantly lower than Black and Hispanic/Latino women. South Asian women had the highest AHEI score (68.5) of all groups (White: 51.5, Chinese: 58.6, Black: 47.7, Hispanic/Latino: 50.3), the lowest alcohol use, and the most exercise.

### Trends Across Age Among Women

South Asian women had a significantly smaller increase in HTN prevalence (4.7%) from age 45 to 55 compared to Chinese (17.2%, p < 0.01), Black (31.0%, p < 0.001), and Latina (14.9%, p < 0.01) women. South Asian women had larger increases in BMI (2.0) compared to White (1.0, p = 0.018) and Chinese (-0.2, p < 0.001) women. White women had a larger increase in AHEI diet score (7.0) compared to South Asian women (3.4, p = 0.006). In addition, Black women had a larger increase in exercise (962 MET min/wk) than South Asian women (203 MET min/wk).

## DISCUSSION

### Principal Findings

In this longitudinal multi-race and ethnic population-based cohort study of 2,700 adults ages 45-55 at baseline in the U.S., we observed significant differences in the prevalence of risk factors for South Asians compared to other U.S. race and ethnic groups at age 45 years. At age 45, South Asian adults had a higher prevalence of prediabetes, diabetes, and hypertension than White adults. Namely, the prevalence of pre-diabetes and diabetes among South Asian men and women is almost twice the prevalence of Black and White individuals. Despite observing a higher prevalence of clinical risk factors among South Asian adults at 45 years, we also found that South Asian adults had the highest (i.e. best) diet quality of all race and ethnic groups, as well as lower alcohol use and higher exercise levels than most other groups. Interestingly, these findings differ from past research showing that South Asians are less likely to meet physical activity recommendations and eat less healthful diets.^18^ However, prior studies included participants across the adult lifespan, and the present analysis only included South Asian adults during middle-age who we found to have more cardioprotective behaviors.

### Secondary Findings

Overall, it appears that as age increases, the prevalence of clinical risk factors increases significantly among men and women. However, as age increased, we also observed a favorable increase in diet quality among all groups and exercise in all groups except for South Asian women. These findings indicate that the increase in ASCVD risk factors among South Asians as they age is in alignment with other groups, but that South Asians may develop some risk factors earlier than other groups. In addition, while health behaviors improved with age among White, Black, Hispanic/Latino, and Chinese adults, the same level of improvement was not seen among South Asian adults.

There are multiple factors that could be contributing to this trend. For example, there may be social and cultural underpinnings to differences in health behaviors as individuals age. The majority of middle- and older-age South Asians in the US are immigrants, and immigration and acculturation have been shown to impact diet and physical activity ^19–21^. Language and cultural barriers may also contribute to reduced health literacy among South Asians living in the U.S. and preventive health messaging reaching other race and ethnic groups may not be reaching South Asians effectively^22^. In addition, clinical ASCVD risk factors are chronic health conditions which are associated with disability and functional decline. The increased burden of ASCVD risk factors among South Asians in middle-age compared to other groups may be impacting their ability to engage in positive health behaviors like increased physical activity.

Previous studies have shown that the number of positive health behaviors at middle-age is associated with less frailty at older ages^23^. In addition, increasing the number of positive health behaviors a person engages in during middle-age is associated with a lower risk of frailty^23^. Given this finding, strategies to enhance healthy behaviors among aging South Asian adults may be needed to reduce disparities. Future studies should replicate this study design in younger populations to determine the prevalence of clinical and behavioral cardiovascular risk factors prior to middle-age and how behavioral and clinical risk factors may change over time to influence ASCVD outcomes. It is possible that health behaviors at younger ages are more predictive of health status at older ages compared to behaviors at middle-age.

### Implications

This is the first study, to our knowledge, to show that the increased burden of ASCVD in South Asians may be attributed to a higher prevalence of clinical risk factors at younger ages. The results of this study indicate that screening and prevention of ASCVD risk must begin early for people of South Asian background given the increased burden of risk factors at younger ages. Despite having the highest diet quality of all the groups included in this study and high levels of exercise at younger ages, South Asians still had a high prevalence of ASCVD risk factors at young ages, especially diabetes and hypertension. Determining the age at which risk factors screening, prevention strategies, and treatment can optimize health outcomes for South Asian adults is fundamental to addressing their increased ASCVD risk.

### Limitations

In this study, participants had to be free of cardiovascular disease when they were enrolled in the study, so this data is potentially excluding individuals who may have early ASCVD and could add to the burden of risk factors. Also, whereas MASALA and MESA offer longitudinal information regarding patient populations that have been traditionally underrepresented in research, the participants in these studies are likely not fully representative of all people of that race or ethnic group living in the United States. For example, MASALA only recruited from two study sites, all of which are large urban areas, excluding many individuals who may add more diversity to the data collected. In addition, longitudinal data heavily relies on patient follow up. Participants with higher educational attainment, have higher socioeconomic status, and have higher health motivation will be more likely to follow up^24^. It is also likely that differences between participants in MESA and MASALA are confounded by time period-based differences, as there is a decade difference between the baseline exams of both studies.

### Conclusions

This study suggests that South Asians have a higher prevalence of several clinical risk factors at age 45, while having more favorable cardiovascular health behaviors, compared to other race and ethnic groups. Given the relatively high observed burden of clinical risk factors in South Asians, future studies should examine the development of risk factors among younger South Asians to better characterize the development and burden of risk earlier in the life course. Given that heterogeneity in risk factors already existed at age 45, future studies should aim to examine the entire life course and identify critical time periods where the burden of risk factors stratifies.

Further research is needed to identify mechanisms of the higher observed burden of clinical risk factors among U.S. South Asians at younger ages despite more favorable cardiovascular health behaviors. Understanding trends and disparities in risk and protective factors across the life course can help equitably improve prevention and treatment strategies for US populations.

## Data Availability

The data used in this study is publically available upon request from the MASALA and MESA research teams.

## ACKNOWLEDGEMENTS

The authors thank the other investigators, the staff, and the participants of the MESA and MASALA studies for their valuable contributions.

## Human Subjects Statement

The Multi-Ethnic Study of Atherosclerosis (MESA) was approved by the institutional review boards at each of the study sites.

## APPENDIX: STATISTICAL METHODS

### A CONTINUOUS RISK FACTORS

#### A.1 MODEL

Let *y*_*ij*_ be the risk factor for participant *i* (*i* = 1, …, *n*) measured at exam *j* (*j* = 1, …, *J*_*i*_). For continuous risk factors (BMI, diet score, physical activity) we fit the following linear mixed-effects model:

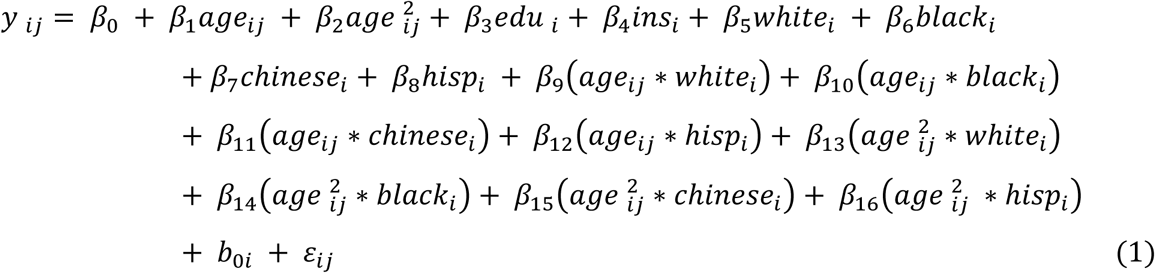

where *age*_*ij*_ is the age of participant *i* at exam *j*, *edu*_*i*_ is a vector of indicator variables for the education categories in Table 1, *ins*_*i*_ is a vector of indicator variables for the insurance categories in Table 1, and *white*_*i*_, *black*_*i*_, *chinese*_*i*_, and *hisp*_*i*_ are indicator variables for race (the reference race is South Asians). The model also includes age by race interaction terms as well as age squared by race interaction terms. The parameters *b*_0*i*_ and ε_*ij*_ are independent random subject-specific intercept and error terms each normally distributed with variance 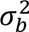 and, 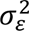 respectively.

#### A.2 PREDICTIVE MARIGINAL MEANS

To calculate the risk factor means by age and race controlling for the effects of education and insurance status, we use the method of predictive marginal means [1]. For example, to obtain the mean risk factor level at age 45 for whites, we set *age*_*ij*_ = 45, 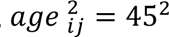 and *white_i_* = 1 and perform the following calculation, using the estimated coefficients (β’*s*) from the model in (1) and the empirical Bayes estimates of the random effects 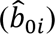. That is,

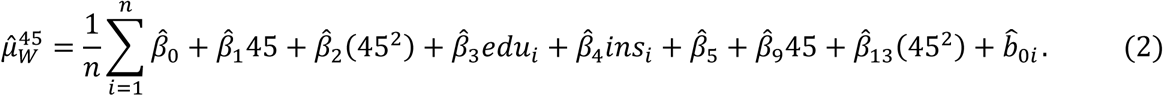

Similar calculations can be performed to obtain predicted means for the other race/ethnic groups at age 45 and age 55.

### B BINARY RISK FACTORS

#### B.1 MODEL

The model for binary risk factors (hypertension, dyslipidemia, diabetes, alcohol use) is similar to that in (1) except we now use a mixed-effects logistic regression model of the probability (*p*_*ij*_) that participant *i* has the risk factor at exam *j*. That is,

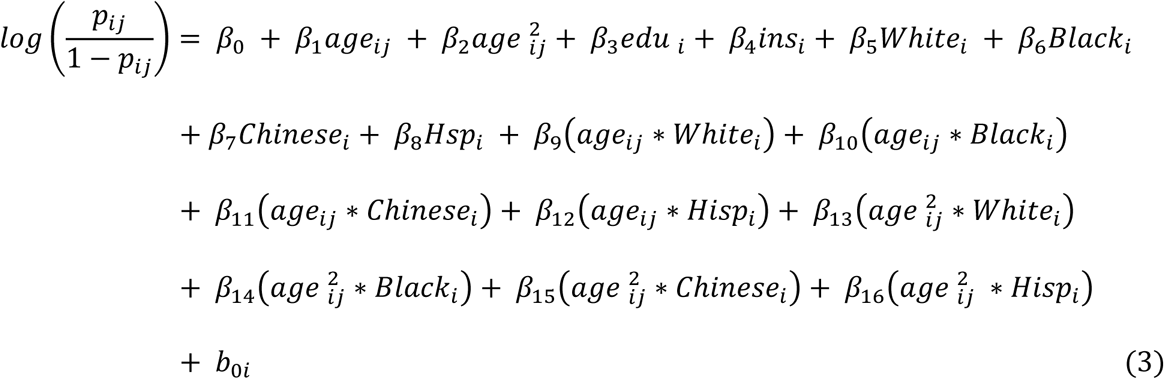

where as in (1) the parameter *b*_0*i*_ is a random intercept term for participant *i*.

#### B.2 PREDICTIVE MARIGINAL MEANS (PROBABILITIES)

Predictive marginal means for the model in (3) can be calculated similar to that in (2). Again, using the example of a White participant at age 45 we calculate the following quantity:

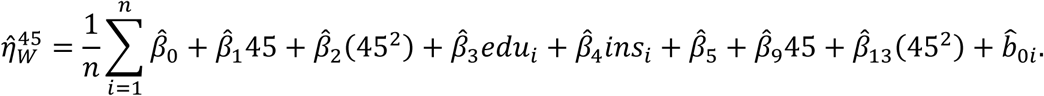

which is the log odds that a white participant has the risk factor at age 45. To put it on the probability scale, we apply the inverse logit transformation:

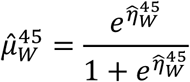

### C CONTRASTS

#### C.1 CONTRASTS AT AGE 45

The first set of contrasts compare predicted marginal mean outcomes at age 45 of White (W), Chinese (C), Black (B), and Hispanics/Latino (H) versus South Asians (SA). We first performed an overall (Wald) test of whether any of these contrasts were significant. That is

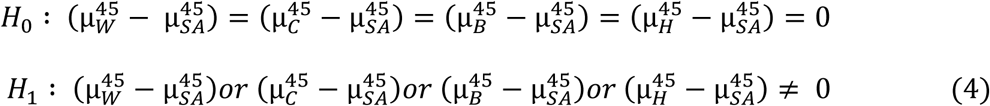

Only if the overall test in (4) is rejected did we perform the four separate comparisons.

#### C.1 SLOPE CONTRASTS

The second set of contrasts compares the change in risk factor levels from age 45 to 55 in South Asians versus the four other race/ethnic groups. Let Δ_*R*_ be the change in predicted risk factor means from age 45 to age 55 for race/ethnic group *R*. That is

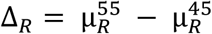

As with the baseline contrasts, we first performed an overall (Wald) test of whether any of the slopes in White, Chinese, Black, and Latino people were significantly different from the South Asian slope, that is

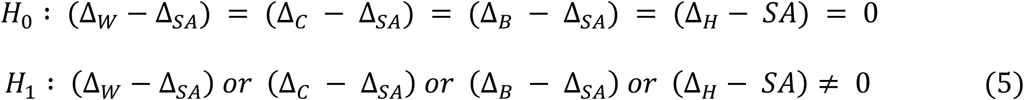

Only if the overall test in (5) is rejected did we perform the four separate tests comparing the rate of change in the four MESA race/ethnic groups to South Asians.

## Sources Cited

1. Prasad DS, Kabir Z, Dash AK and Das BC. Childhood cardiovascular risk factors in South Asians: A cause of concern for adult cardiovascular disease epidemic. Ann Pediatr Cardiol. 2011;4:166–71.

2. Stanford South Asian Translational Heart Initiative.

3. Gupta K, Modi S and Ananthasubramaniam K. Toward Understanding Cardiovascular Risk Burden in South Asians: A Major Step Forward. JACC Asia. 2022;2:912–915.

4. Kanaya AM, Kandula N, Herrington D, Budoff MJ, Hulley S, Vittinghoff E and Liu K. Mediators of Atherosclerosis in South Asians Living in America (MASALA) study: objectives, methods, and cohort description. Clin Cardiol. 2013;36:713–720.

5. Bild DE, Bluemke DA, Burke GL, Detrano R, Diez Roux AV, Folsom AR, Greenland P, Jacob DR, Jr., Kronmal R, Liu K, Nelson JC, O’Leary D, Saad MF, Shea S, Szklo M and Tracy RP. Multi-Ethnic Study of Atherosclerosis: objectives and design. Am J Epidemiol. 2002;156:871–81.

6. Lloyd-Jones DM, Allen NB, Anderson CAM, Black T, Brewer LC, Foraker RE, Grandner MA, Lavretsky H, Perak AM, Sharma G, Rosamond W and American Heart A. Life’s Essential 8: Updating and Enhancing the American Heart Association’s Construct of Cardiovascular Health: A Presidential Advisory From the American Heart Association. Circulation. 2022;146:e18–e43.

7. Kanaya AM, Herrington D, Vittinghoff E, Ewing SK, Liu K, Blaha MJ, Dave SS, Qureshi F and Kandula NR. Understanding the high prevalence of diabetes in U.S. south Asians compared with four racial/ethnic groups: the MASALA and MESA studies. Diabetes Care. 2014;37:1621–8.

8. Gujral UP, Vittinghoff E, Mongraw-Chaffin M, Vaidya D, Kandula NR, Allison M, Carr J, Liu K, Narayan KMV and Kanaya AM. Cardiometabolic Abnormalities Among Normal-Weight Persons From Five Racial/Ethnic Groups in the United States: A Cross-sectional Analysis of Two Cohort Studies. Ann Intern Med. 2017;166:628–636.

9. Rodriguez LA, Jin Y, Talegawkar SA, Otto MCO, Kandula NR, Herrington DM and Kanaya AM. Differences in Diet Quality among Multiple US Racial/Ethnic Groups from the Mediators of Atherosclerosis in South Asians Living in America (MASALA) Study and the Multi-Ethnic Study of Atherosclerosis (MESA). J Nutr. 2020;150:1509–1515.

10. Chobanian AV, Bakris GL, Black HR, Cushman WC, Green LA, Izzo JL, Jr., Jones DW, Materson BJ, Oparil S, Wright JT, Jr., Roccella EJ, Joint National Committee on Prevention DE, Treatment of High Blood Pressure. National Heart L, Blood I and National High Blood Pressure Education Program Coordinating C. Seventh report of the Joint National Committee on Prevention, Detection, Evaluation, and Treatment of High Blood Pressure. Hypertension. 2003;42:1206–52.

11. Genuth S, Alberti KG, Bennett P, Buse J, Defronzo R, Kahn R, Kitzmiller J, Knowler WC, Lebovitz H, Lernmark A, Nathan D, Palmer J, Rizza R, Saudek C, Shaw J, Steffes M, Stern M, Tuomilehto J, Zimmet P, Expert Committee on the D and Classification of Diabetes M. Follow-up report on the diagnosis of diabetes mellitus. Diabetes Care. 2003;26:3160–7.

12. Expert Panel on Detection E and Treatment of High Blood Cholesterol in A. Executive Summary of The Third Report of The National Cholesterol Education Program (NCEP) Expert Panel on Detection, Evaluation, And Treatment of High Blood Cholesterol In Adults (Adult Treatment Panel III). JAMA. 2001;285:2486–97.

13. Consultation WHOE. Appropriate body-mass index for Asian populations and its implications for policy and intervention strategies. Lancet. 2004;363:157–63.

14. Defining Adult Overweight & Obesity.

15. Ainsworth BE, Irwin ML, Addy CL, Whitt MC and Stolarczyk LM. Moderate physical activity patterns of minority women: the Cross-Cultural Activity Participation Study. J Womens Health Gend Based Med. 1999;8:805–13.

16. Williams R. Using the margins command to estimate and interpret adjusted predictions and marginal effects. The Stata Journal. 2012;12:308–331.

17. Stata 18 Base Reference Manual [computer program]. College Station, TX: Stata Press; 2023.

18. Nair MP, D. Why Do South Asians Have High Risk for CAD? Global Heart 2012;7:307–314.

19. Patel M, Phillips-Caesar E and Boutin-Foster C. Barriers to lifestyle behavioral change in migrant South Asian populations. J Immigr Minor Health. 2012;14:774–85.

20. Zhu X, Liu J, Sevoyan M and Pate RR. Acculturation and leisure-time physical activity among Asian American adults in the United States. Ethn Health. 2022;27:1900–1914.

21. Dave SS, Craft LL, Mehta P, Naval S, Kumar S and Kandula NR. Life stage influences on U.S. South Asian women’s physical activity. Am J Health Promot. 2015;29:e100–8.

22. Maresova P, Javanmardi E, Barakovic S, Barakovic Husic J, Tomsone S, Krejcar O and Kuca K. Consequences of chronic diseases and other limitations associated with old age - a scoping review. BMC Public Health. 2019;19:1431.

23. Gil-Salcedo A, Dugravot A, Fayosse A, Dumurgier J, Bouillon K, Schnitzler A, Kivimaki M, Singh-Manoux A and Sabia S. Healthy behaviors at age 50 years and frailty at older ages in a 20-year follow-up of the UK Whitehall II cohort: A longitudinal study. PLoS Med. 2020;17:e1003147.

24. Guo A, Jin H, Mao J, Zhu W, Zhou Y, Ge X and Yu D. Impact of health literacy and social support on medication adherence in patients with hypertension: a cross-sectional community-based study. BMC Cardiovasc Disord. 2023;23:93.

